# Multimodal EHR-Based Prediction of Pediatric Asthma Exacerbations

**DOI:** 10.64898/2026.02.25.26347091

**Authors:** Zhengkang Fan, Jinqian Pan, Mengxian Lyu, Renjie liang, Chengkun Sun, Yonghui Wu, David Fedele, Jennifer Fishe, Jie Xu

## Abstract

Pediatric asthma exacerbations are a frequent cause of emergency department (ED) visits and hospitalizations, yet accurate risk prediction remains limited and no consensus risk scores exist. Using UF Health electronic health records (EHRs) from 2011-2023, we evaluated two computable phenotypes (i.e., CAPriCORN and COMPAC) to predict exacerbations over 6-, 12-, and 24-month horizons. Exacerbations were defined using a validated composite of diagnosis codes from ED, inpatient, or outpatient encounters combined with systemic corticosteroids prescriptions. Several commonly used machine learning (ML) models were trained with stratified five-fold cross-validation, Bayesian hyperparameter optimization, and Youden’s J thresholding. XGBoost achieved the best performance, with SHapley Additive exPlanations (SHAP) highlighting note-derived symptom terms and rescue-medication use as dominant predictors. Future work will focus on external validation and assessment of generalizability. This interpretable, text-integrated framework may support child-specific risk stratification and inform EHR-based decision support for timely pediatric asthma management.

## 1 Introduction

Childhood asthma is a highly prevalent chronic disease in the United States, with approximately 4.7 million children under 18 years living with asthma in 2021. Exacerbations—acute worsening of respiratory symptoms—are common, with 38 7% of children with asthma reporting at least one exacerbation in the previous year. ^1,2^ Exacerbations also drive acute healthcare use: in 2020, there were an estimated 270,330 pediatric emergency department (ED) visits and 27,055 hospital inpatient stays nationally for asthma. ^2^ Hospitalizations impose measurable economic burden; for example, potentially preventable pediatric asthma inpatient stays accounted for $278.1 million in aggregate hospital costs in 2017 (mean $5,200 per stay). ^3^ Despite this burden, early prediction of asthma exacerbations in children remains challenging, and no consensus clinical prediction tools are widely used. Recent reviews highlight key research gaps, including data quality, generalizability, and clinical integration of predictive approaches. ^4–6^

Most existing studies leverate electronic health record (EHR) data—including diagnoses, encounters, vitals, labs, and medications—with multiple machine learning (ML) models demonstrating good performance for exacerbation risk estimation. However, many models are derived from adult or mixed-age cohorts, limiting applicability to pediatrics, where pathophysiology, triggers, and care pathways differ. ^7,8^ For example, Xiang et al. built a time-sensitive attentive neural network on longitudinal EHR to forecast adult exacerbations; ^9^ Lisspers et al. trained ML models on Swedish primary care EHR for short-term prediction; ^10^ and Zein et al. applied modern ML to tertiary care EHR to identify adults at risk. ^11^

Pediatric-focused exacerbation prediction research often relies on a single modality or a limited set of structured EHR features, which may not capture salient characteristics of a heterogeneous chronic disease that with varied exacerbation triggers. Many studies also focus on predicting hospitalization after ED arrival rather than the exacerbation itself. ^4–6^ For instance, Murugan et al. assessed a commercial risk model using routinely collected EHR fields; ^12^ Sills et al. leveraged ED triage variables to predict hospitalization among children with asthma; ^13^ and AlSaad et al. applied deep learning to EHR data to forecast ED utilization in pediatric asthma. ^14^ Even studies labeled “multimodal” often perform shallow integration, appending a small set of environmental exposures or point-of-care context to structured EHR features rather than performing deep, longitudinal multisource fusion. For example, Hurst et al. combined pediatric EHR data with a limited panel of environmental indicators, while triage-time models such as Patel et al. focus on presentation-time clinical variables and add weather (e.g., temperature, humidity), neighborhood characteristics, and community viral activity to predict hospitalization at ED arrival. ^15,16^ These approaches are further limited by sparse outcome labels, small sample sizes, and site-specific coding variability, weakening generalization across diverse pediatric populations and for dynamic short-term risk prediction.

Recent evidence indicates that the symptom information captured in clinical notes is strongly associated with acute asthma exacerbations. NLP-extracted mentions of wheeze, cough, and related trajectories predict near-term risk in adult cohorts. ^17,18^ Because free text details—such as nocturnal cough, rescue inhaler overuse, viral illness, and clinician impressions—are rarely codified in structured EHR fields, integrating clinical notes represents an opportunity to augment structured signals.

To address this gap, we developed and evaluated ML models that integrate structured EHR features with clinical notes to predict pediatric asthma exacerbations treated across inpatient and outpatient settings. Leveraging a large multimodal pediatric cohort, we operationalized two computable phenotypes (CPs), compared five complementary ML models, and assessed performance across multiple prediction windows to evaluate whether text-integrated models better capture pediatric asthma variability and improve discrimination and timeliness. Clinically, more accurate near-term prediction could enable early, individualized risk stratification to guide proactive outreach and step-up therapy, potentially reducing ED visits and hospitalizations.

## 2 Method

### 2.1 Data source and study population

Data were obtained from the University of Florida (UF) Integrated Data Repository (IDR), a large scale clinical data warehouse that consolidates information across the UF Health system, including the Epic electronic health record (Janesville, WI). The IDR contains longitudinal data for over two million patients, encompassing demographics, encounters, diagnosis codes, medications, and unstructured clinical narratives. The study was approved by the University of Florida Institutional Review Board (IRB#202002779).

To identify pediatric asthma patients, we first extracted records for more than 128,000 children with asthma-related conditions between 2011 and 2023. Two published computable phenotypes (CPs)—CAPriCORN ^19^ and COMPAC ^20^— were applied according to their original rule and value sets. CAPriCORN is a rule-based algorithm using structured EHR elements (asthma diagnosis codes and asthma medication exposures) to define cases. In contrast, COMPAC combines structured criteria (diagnoses, inhaled bronchodilators, corticosteroid prescriptions) with text-derived evidence of asthma-related signs and symptoms extracted from clinical notes using keyword rules. COMPAC was originally developed and validated on the UF Health IDR data. Table 1 summarizes each CP’s performance compared with chart reviewed results. ^19,20^ The study population was limited to children aged 2-18 years, excluding younger children to reduce potential confounding from bronchiolitis-induced wheezing, a common cause of ED visits in that age group.

**Table 1.**
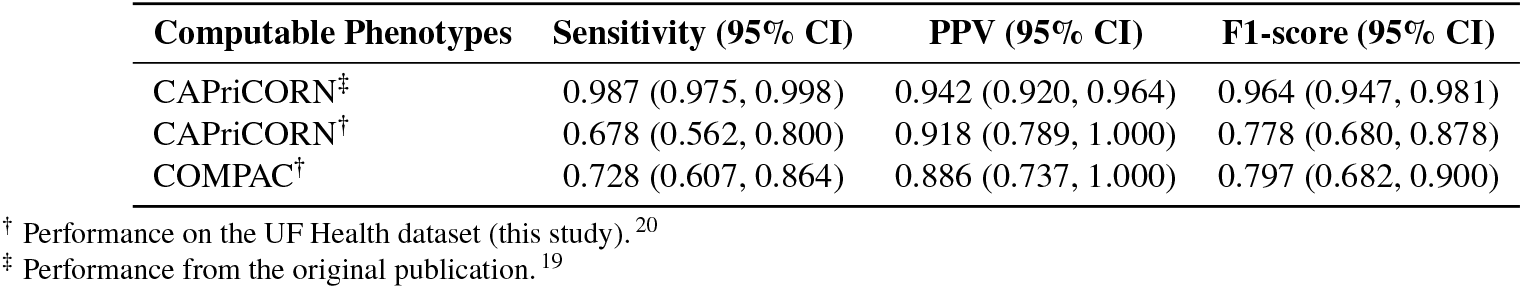
Performance comparison of the Computable Phenotypes (i.e., CAPriCORN and COMPAC).

### 2.2 Definition of asthma exacerbation

Asthma exacerbations were defined in alignment with prior EHR-based studies, harmonizing criteria across care settings. Definitions in the literature vary—some identify events through ED visits with asthma diagnoses; ^21^ others use medication-based proxies, typically short courses of systemic corticosteroids; ^15,16^ still others define events by hospital admissions or ED visits; ^22,23^ and several employ composite endpoints that combine medication exposure with ED and/or inpatient encounters. ^24,25^ Consistent with composite definitions, we classified an encounter as an asthma exacerbation if any of the following criteria were met: (1) an ED visit with an asthma diagnosis; (2) a hospital admission with an asthma diagnosis; or (3) an outpatient visit with an asthma diagnosis accompanied by asthma-related systemic corticosteroid prescription (administered or dispensed) linked to the encounter. Encounters meeting any single criterion were labeled as asthma exacerbations.

### 2.3 Observation period and prediction window

As illustrated in the lower panel of **Fig. 1**, the analytic timeline was anchored at the index date, defined as the first encounter with an asthma diagnosis. The observation period extended from the earliest recorded encounter in UF Health through the index date. Prediction windows of 6 months, 1 year, and 2 years following the index date were then evaluated. For each window, the occurrence of ≥1 asthma exacerbation were labeled as positive (exacerbation risk), while absence indicated negative (no risk). Each window was analyzed independently, allowing patients to have different outcomes across horizons.

**Figure 1.**
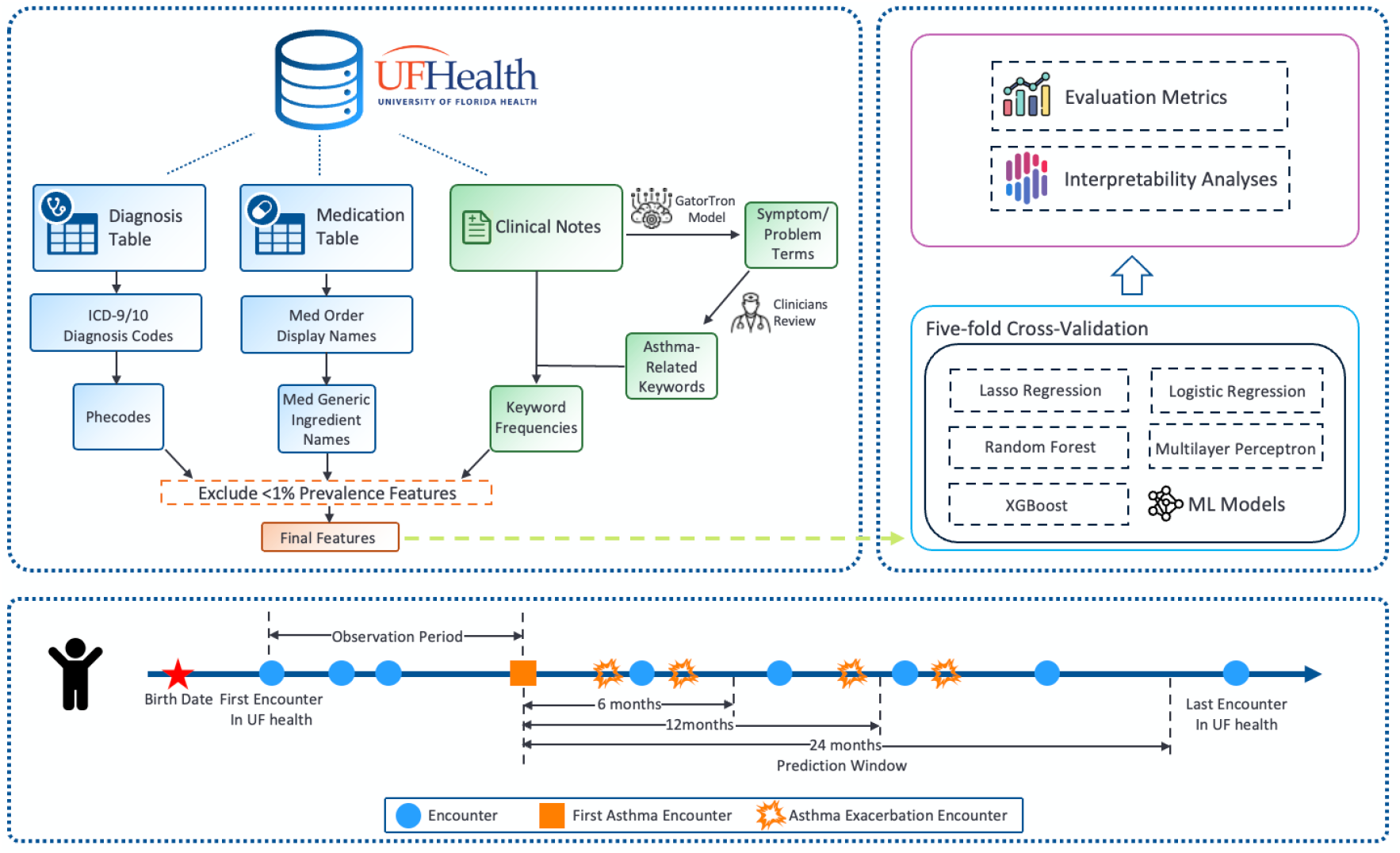
Schematic of the feature engineering process (top) and corresponding patient timeline illustrating observation and prediction windows (bottom).

### 2.4 Feature engineering

During the observation period, features were derived from structured EHR data (comorbidities and medications) and clinical narratives capturing asthma-related terms (see upper panel of Fig. 1). Comorbidities were represented by mapping all ICD-9/10 diagnosis codes to Phecodes to reduce dimensionality and sparsity. ^26^ Medication exposures were aggregated at the ingredient (generic) level rather than product names to consolidate equivalent formulations. For clinical notes,Asthma-related features were extracted from longitudinal clinical notes using a GatorTron-based named entity recognition pipeline restricted to problem entities. GatorTron, ^27^ pretrained on large-scale clinical narratives and fine-tuned on the 2010 i2b2 dataset, was applied to all patient notes to identify span-level problem mentions. Extracted terms were aggregated across the cohort, normalized to canonical forms (e.g., standardizing case, punctuation, and common abbreviations), and ranked by frequency; the 1,000 most frequent candidates as an initial asthma keyword set were retained for review. Two clinicians (JF and DF) then reviewed and merged overlapping or synonymous concepts to minimize redundancy, producing a refined set of asthma-related keywords. Term frequencies of these keywords were computed as note-derived features. Finally, after combining Phecode-mapped diagnoses, ingredient-level medications, and note-derived term frequencies, features with an overall prevalence <1% in the computable-phenotype cohort were excluded to reduce sparsity, yielding the final feature set for modeling.

### 2.5 Machine learning algorithms and experimental settings

Five supervised machine learning models were evaluated: Lasso regression (LASSO), logistic regression (LR), multilayer perceptron (MLP), random forest (RF), and extreme gradient boosting (XGBoost). Models were trained and evaluated using stratified five-fold cross-validation. Hyperparameters were tuned via Bayesian optimization on the first fold, with the optimal configuration applied to the remaining four folds. Performance was primarily assessed by the area under the receiver operating characteristic curve (AUC). For threshold-dependent metrics (accuracy, F1 score, precision, recall, and specificity), predicted probabilities were binarized using the threshold that maximized Youden’s J statistic.

### 2.6 Identification of important predictors

We applied SHAP (SHapley Additive exPlanations) ^28^ to quantify each predictor’s contribution to the model output and to characterize its association with the outcome. For the best-performing models, SHAP bar and summary plots highlighted the top 15 predictors. Positive SHAP values indicate variables increasing the model-predicted risk of exacerbation, while negative values indicate protective factors. Feature importance was ranked by the absolute mean SHAP value across patients.

## 3 Result

### 3.1 Cohort characteristics

As shown in Table 2, the pediatric asthma cohorts identified by the CAPriCORN and COMPAC computable phenotypes demonstrated similar demographic and clinical characteristics. Both cohorts included more than 27,000 children. The sex distribution was nearly identical, with approximately 57% male and 43% female participants. Racial and ethnic composition followed similar patterns, with non-Hispanic Black children representing the largest subgroup (43%), followed by non-Hispanic White children (38%), Hispanic children (10%), and a smaller proportion classified as other.

**Table 2.** Characteristics of the pediatric asthma cohorts identified by CAPriCORN and COMPAC.

| Characteristics | CAPriCORN (N = 27,827) | COMPAC (N = 28,878) |
| --- | --- | --- |
| **Age, N (%)** |  |  |
| 2 ~ 5 | 12,626 (45.4) | 12,708 (44.0) |
| 6 ~ 12 | 9,513 (34.2) | 10,104 (35.0) |
| 13 ~ 18 | 5,164 (18.6) | 5,460 (18.9) |
| **Sex, N (%)** |  |  |
| Male | 15,737 (56.6) | 16,379 (56.7) |
| Female | 12,089 (43.4) | 12,498 (43.3) |
| Unknown | 1 (0.0) | 1 (0.0) |
| **Race/Ethnicity, N (%)** |  |  |
| NHW | 10,431 (37.5) | 10,923 (37.8) |
| NHB | 12,017 (43.2) | 12,382 (42.9) |
| Hispanic | 2,727 (9.8) | 2,843 (9.8) |
| Other | 905 (3.3) | 933 (3.2) |
| **Asthma Exacerbation Risk, N (%)** |  |  |
| In 6 months | 2,915 (10.5) | 2,899 (10.0) |
| In 12 months | 4,249 (15.3) | 4,230 (14.6) |
| In 24 months | 5,637 (20.3) | 5,614 (19.4) |
| **#Exacerbations Per Patient, Mean (SD)** |  |  |
| In 6 months | 1.38 (0.85) | 1.39 (0.85) |
| In 12 months | 1.64 (1.23) | 1.64 (1.23) |
| In 24 months | 1.98 (1.79) | 1.99 (1.80) |

Asthma exacerbation rates were slightly higher in the CAPriCORN cohort compared to COMPAC, with differences of approximately 0.5% to 1% across the examined follow-up periods. Overall, the proportion of patients experiencing at least one exacerbation was around 10% within 6 months of the index date, 15% within 1 year, and 20% within 2 years.

### 3.2 Modeling results

As shown in Table 3, machine learning models achieved stable performance across all prediction windows, with XG-Boost consistently outperforming other approaches. In the 6-month prediction window, XGBoost achieved the highest AUC values in both cohorts (0.813 for COMPAC and 0.827 for CAPriCORN). Similar trends were observed at 12 months, where XGBoost again yielded superior performance (AUC 0.844 for COMPAC and 0.813 for CAPriCORN). At 24 months, XGBoost maintained its advantage, reaching AUCs of 0.829 in COMPAC and 0.848 in CAPriCORN, the highest across all models and time horizons. Although LR, MLP, and RF models demonstrated competitive performance, their predictive ability remained consistently lower than that of XGBoost. These findings highlight the robustness of XGBoost for predicting pediatric asthma exacerbations across different time horizons and computable phenotypes.

**Table 3.** Model performance for predicting asthma exacerbations across prediction windows, CPs, and machine learning models (mean±SD). The best AUC for each CP and window is bolded.

| Prediction Window | CP | Model | Accuracy | F1 | Precision | Recall | AUC | Specificity |
| --- | --- | --- | --- | --- | --- | --- | --- | --- |
| In 6 months | COMPAC | LASSO | 0.607±0.055 | 0.684±0.044 | 0.865±0.004 | 0.607±0.055 | 0.678±0.013 | 0.600±0.068 |
| | | LR | 0.715±0.031 | 0.770±0.023 | 0.884±0.002 | 0.715±0.031 | 0.777±0.006 | 0.715±0.037 |
| | | MLP | 0.713±0.034 | 0.769±0.025 | 0.889±0.002 | 0.713±0.034 | 0.795±0.008 | 0.708±0.041 |
| | | RF | 0.707±0.018 | 0.764±0.014 | 0.885±0.004 | 0.707±0.018 | 0.779±0.011 | 0.704±0.024 |
| | | XGBoost | 0.739±0.023 | 0.788±0.017 | 0.890±0.002 | 0.739±0.023 | **0.813±0.006** | 0.738±0.028 |
| | CAPriCORN | LASSO | 0.651±0.024 | 0.702±0.019 | 0.827±0.002 | 0.651±0.024 | 0.720±0.005 | 0.643±0.033 |
| | | LR | 0.716±0.017 | 0.755±0.013 | 0.847±0.002 | 0.716±0.017 | 0.791±0.011 | 0.713±0.024 |
| | | MLP | 0.732±0.021 | 0.768±0.016 | 0.853±0.002 | 0.732±0.021 | 0.810±0.006 | 0.728±0.029 |
| | | RF | 0.728±0.025 | 0.765±0.020 | 0.848±0.003 | 0.728±0.025 | 0.793±0.007 | 0.729±0.034 |
| | | XGBoost | 0.739±0.030 | 0.774±0.023 | 0.859±0.003 | 0.739±0.030 | **0.827±0.005** | 0.734±0.040 |
| In 12 months | COMPAC | LASSO | 0.664±0.009 | 0.698±0.008 | 0.797±0.002 | 0.664±0.009 | 0.747±0.012 | 0.647±0.013 |
| | | LR | 0.724±0.011 | 0.750±0.009 | 0.821±0.002 | 0.724±0.011 | 0.810±0.007 | 0.715±0.016 |
| | | MLP | 0.732±0.023 | 0.758±0.019 | 0.829±0.002 | 0.732±0.023 | 0.828±0.007 | 0.722±0.034 |
| | | RF | 0.728±0.028 | 0.753±0.023 | 0.818±0.005 | 0.728±0.028 | 0.809±0.007 | 0.725±0.043 |
| | | XGBoost | 0.752±0.025 | 0.774±0.020 | 0.835±0.002 | 0.752±0.025 | **0.844±0.003** | 0.746±0.040 |
| | CAPriCORN | LASSO | 0.580±0.061 | 0.658±0.056 | 0.861±0.004 | 0.580±0.061 | 0.680±0.017 | 0.565±0.075 |
| | | LR | 0.724±0.022 | 0.775±0.016 | 0.880±0.004 | 0.724±0.022 | 0.778±0.011 | 0.725±0.028 |
| | | MLP | 0.717±0.016 | 0.771±0.012 | 0.886±0.002 | 0.717±0.016 | 0.797±0.008 | 0.712±0.020 |
| | | RF | 0.722±0.025 | 0.774±0.019 | 0.888±0.002 | 0.722±0.025 | 0.804±0.009 | 0.716±0.029 |
| | | XGBoost | 0.719±0.038 | 0.772±0.029 | 0.890±0.002 | 0.719±0.038 | **0.813±0.009** | 0.712±0.047 |
| In 24 months | COMPAC | LASSO | 0.636±0.037 | 0.686±0.032 | 0.823±0.003 | 0.636±0.037 | 0.724±0.011 | 0.621±0.050 |
| | | LR | 0.729±0.029 | 0.763±0.022 | 0.835±0.003 | 0.729±0.029 | 0.793±0.005 | 0.731±0.042 |
| | | MLP | 0.719±0.035 | 0.756±0.028 | 0.850±0.003 | 0.719±0.035 | 0.812±0.005 | 0.711±0.050 |
| | | RF | 0.741±0.020 | 0.772±0.016 | 0.843±0.002 | 0.741±0.020 | 0.796±0.009 | 0.746±0.027 |
| | | XGBoost | 0.751±0.026 | 0.782±0.021 | 0.845±0.002 | 0.751±0.026 | **0.829±0.008** | 0.751±0.050 |
| | CAPriCORN | LASSO | 0.669±0.027 | 0.700±0.024 | 0.794±0.002 | 0.669±0.027 | 0.750±0.006 | 0.653±0.046 |
| | | LR | 0.715±0.018 | 0.741±0.016 | 0.817±0.002 | 0.715±0.018 | 0.811±0.011 | 0.699±0.027 |
| | | MLP | 0.734±0.020 | 0.757±0.016 | 0.823±0.002 | 0.734±0.020 | 0.828±0.010 | 0.724±0.033 |
| | | RF | 0.727±0.016 | 0.751±0.013 | 0.817±0.003 | 0.727±0.016 | 0.813±0.008 | 0.720±0.026 |
| | | XGBoost | 0.762±0.020 | 0.781±0.016 | 0.831±0.003 | 0.762±0.020 | **0.848±0.006** | 0.760±0.033 |

### 3.3 Interpretability analyses

SHAP analyses indicated that, for both CPs, note-derived respiratory symptoms were the most informative features across all prediction windows. These include keywords indicating dyspnea on exertion, asthma, wheeze, and cough. Medication prescriptions or administrations—particularly the rescue medicine albuterol sulfate and the long-term medication antihistamine cetirizine—as well as allergy-related diagnoses also showed substantial importance. Physical examination findings, such as accessory muscle use and nasal flaring, contributed additional predictive signals. The ranking of predictors remained broadly stable from 6 to 24 months. CAPriCORN placed relatively greater weight on infection or other respiratory disease phenotypes at longer horizons, whereas COMPAC emphasized note-based symptoms and short-acting bronchodilator use. Positive SHAP values corresponded to increased risk, supporting the face validity of both models and highlighting the dominant role of free text symptom keywords and rescue medication use in pediatric asthma risk stratification (Fig. 2).

**Figure 2.**
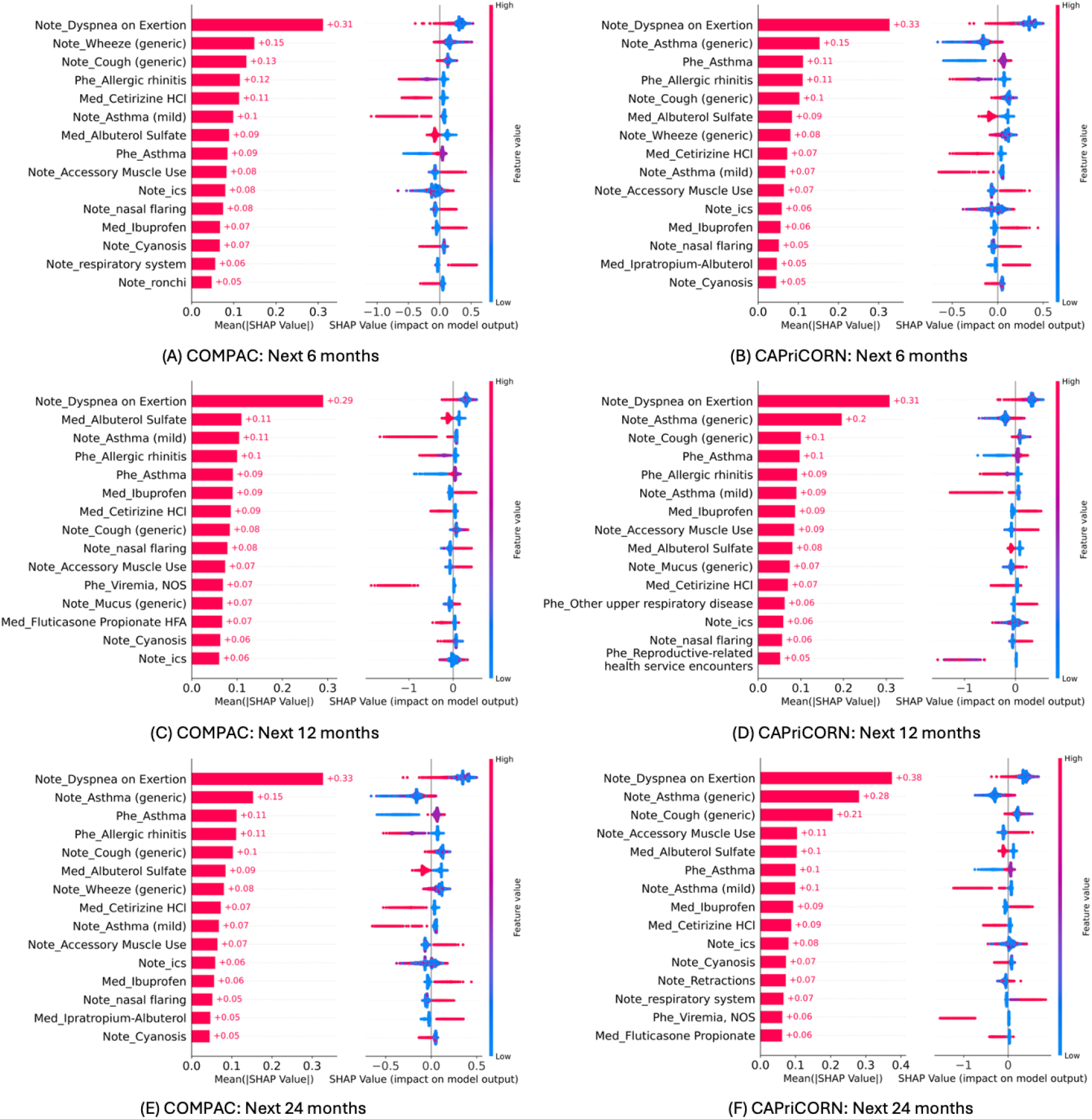
SHAP-derived feature importance for COMPAC and CAPriCORN across 6-, 12-, and 24-month prediction windows.

## 4 Discussion

### 4.1 Predictive performance of machine learning models

In this study, ML models combining structured EHR features with NLP-derived information from clinical notes achieved consistent performance in predicting pediatric asthma exacerbations at 6, 12, and 24 months. XGBoost consistently outperformed other classifiers across both CP cohorts (Table 3), achieving AUCs of 0.813 (COMPAC) and 0.827 (CAPriCORN) at 6 months; 0.844 (COMPAC) and 0.813 (CAPriCORN) at 12 months; 0.829 (COMPAC) and 0.848 (CAPriCORN) at 24 months. These results align with prior evidence showing that richer, multi-modal clinical representations improve predictive performance, whereas previous pediatric EHR-based studies often relied on single modality inputs, ^12–14^ shallow text integration, ^15,16^ and showed limited generalizability. ^7,8^

By integrating GatorTron-derived symptom terms from clinical notes with structured diagnoses and medications, our approach demonstrates the potential of multi-modal modeling while exploring portability across pediatric cohorts and prediction windows. The consistent performance of XGBoost suggests that gradient-boosted trees offer a robust approach for multi-horizon risk forecasting in pediatric populations.

### 4.2 Interpretability and clinical implications

SHAP analyses indicated that note-derived respiratory symptoms (e.g., dyspnea on exertion, wheeze, and cough), recent rescue-medication exposure (i.e., albuterol), and allergy diagnoses were the most influential predictors, with rankings remaining stable across time horizons (Fig. 2). These findings align with prior studies suggesting that NLP-extracted symptom trajectories, especially repeated wheeze and dyspnea, can help forecast near-term exacerbation events. ^17^ Additionally, the prominence of allergic comorbidities is consistent with the well-recognized link between allergies and asthma exacerbation risk, where effective management of rhinitis may help reduce exacerbation events in children. ^29,30^

Two medications-related observations merit cautious interpretation. First, ibuprofen exposure was associated with higher predicted risk across cohorts and horizons. Evidence in pediatric asthma literature is mixed: randomized trials generally not shown increased exacerbation risk with as-needed ibuprofen use, ^31,32^ through observational studies highlight potential concerns, particularly for children with NSAID-exacerbated respiratory disease. Ibuprofen use may also indicate intercurrent viral illness, which itself can trigger an exacerbation. ^33^ Further analyses are needed to clarify whether the association reflects underlying illness severity, confounding by indication, or a subgroup effect. Second, cetirizine exposure was associated with lower predicted risk, consistent with literature suggesting that treatment of allergic rhinitis can modestly reduce asthma symptoms and exacerbation frequency in sensitized children. ^34^ While this may reflect a true protective association, it may also capture patterns of regular allergy management rather than a direct treatment effect.

### 4.3 Limitations and future directions

This analysis was conducted retrospectively within a single health system, which limits causal inference and may introduce spectrum and documentation biases. The composite EHR-defined outcome, while practical for large-scale analysis, may be vulnerable to coding variability, incomplete capture of out-of-network events, and heterogeneous note quality. Feature construction relied on routinely collected data, and residual confounding cannot be excluded. External generalizability has not yet been established, and environmental factors were not included. Medication-specific associations should be interpreted cautiously and require prospective validation.

Future work should include external and multi-site validation with harmonized feature definitions (e.g., crosswalks for diagnoses and medications) to assess portability. Subgroup fairness and calibration analyses (by age, sex, race/ethnicity, insurance, and baseline severity) should also be performed, with recalibration or group-specific thresholds applied if needed. Methodologically, temporally aware modeling approaches (e.g., visit-level sequences, time-varying covariates, and drift detection) could help capture evolving risk. Systematic error analysis could further characterize false positives and negatives and identify opportunities for model improvement. For free-text note data, robustness to documentation variability (note density, specialty mix) should be tested, and portability across different EHR systems—ideally via standardized data models and containerized pipelines—should be evaluated to facilitate reproducible deployment.

From a clinical perspective, SHAP analyses may help guide the development of EHR- and mHealth-embedded risk scores and alerts, and support interpretability for clinicians and caregivers. While XGBoost is less inherently interpretable than simpler models, user-centered design could provide explanations of how predicted risk changes and which features contribute most. After external validation, such models could inform pre-emptive step-up therapy, reinforce asthma action plans, and support targeted outreach to at-risk patients, with the potential to reduce ED visits, hospitalizations, symptom duration, and missed school days. ^35,36^ To further evaluate potential clinical utility, decision curve analysis and workflow-based implementation studies (silent-mode followed by interventional trials) should be conducted.

## 5 Conclusion

ML models combining structured EHR data with NLP-derived symptom terms achieved consistent, clinically plausible prediction of pediatric asthma exacerbations over 6-, 12-, and 24-month horizons across two CP cohorts (Table 3). Feature attribution highlighted note-derived respiratory symptoms, recent rescue medication exposure, and allergy diagnoses as stable, high-importance predictors (Fig. 2), supporting individualized, multi-horizon risk stratification. These results support the potential development of an EHR-embedded, continuously updated risk score to inform guideline-based step-up therapy, reinforce asthma action plans, and enable targeted outreach. Given the single-system, retrospective design and potential outcome misclassification, external validation, fairness and calibration assessments, and prospective workflow studies are required to assess generalizability and clinical utility. This work provides a practical framework to move from accurate risk prediction toward measurable reductions in preventable pediatric asthma exacerbations and associated healthcare utilization.

## Data Availability

All data produced in the present study are available upon reasonable request to the authors

## Acknowledge

This study was supported by the National Institutes of Health/National Heart, Lung, and Blood Institute (R01HL169277). The content is solely the responsibility of the authors and does not necessarily represent the official views of the National Institutes of Health.

## References

1. Pate CA, Zahran HS. The Status of Asthma in the United States. Preventing Chronic Disease. 2024;21:240005. Available from: https://www.cdc.gov/pcd/issues/2024/24_0005.htm.

2. Centers for Disease Control and Prevention. Most Recent National Asthma Data; 2024. Accessed 2025-09-11. https://www.cdc.gov/asthma/most_recent_national_asthma_data.htm. Available from: https://www.cdc.gov/asthma/most_recent_national_asthma_data.htm.

3. McDermott KW, Jiang HJ. Characteristics and Costs of Potentially Preventable Inpatient Stays, 2017. Rockville, MD: Agency for Healthcare Research and Quality (AHRQ); 2020. 259. Available from: https://hcup-us.ahrq.gov/reports/statbriefs/sb259-Potentially-Preventable-Hospitalizations-2017.jsp.

4. Zhou C, Shuai L, Hu H, Ung COL, Lai Y, Fan L, et al. Applications of Machine Learning Approaches for Pediatric Asthma Exacerbation Management: A Systematic Review. BMC Medical Informatics and Decision Making. 2025;25(1):170. Available from: 10.1186/s12911-025-02990-0.

5. Ojha T, Patel A, Sivapragasam K, Sharma R, Vosoughi T, Skidmore B, et al. Exploring Machine Learning Applications in Pediatric Asthma Management: Scoping Review. JMIR AI. 2024;3:e57983. Available from: https://ai.jmir.org/2024/1/e57983.

6. Votto M, De Silvestri A, Postiglione L, De Filippo M, Manti S, La Grutta S, et al. Predicting Paediatric Asthma Exacerbations with Machine Learning: A Systematic Review with Meta–Analysis. European Respiratory Review: An Official Journal of the European Respiratory Society. 2024;33(174):240118.

7. Budiarto A, Tsang K, Wilson A, Sheikh A, Shah S. Machine Learning–Based Asthma Attack Prediction Models From Routinely Collected Electronic Health Records: Systematic Scoping Review. JMIR AI. 2023;2:e46717. Available from: https://ai.jmir.org/2023/1/e46717.

8. Molfino NA, Turcatel G, Riskin D. Machine Learning Approaches to Predict Asthma Exacerbations: A Narrative Review. Advances in Therapy. 2024;41:534–52.

9. Xiang Y, Ji H, Zhou Y, Li F, Du J, Rasmy L, et al. Asthma Exacerbation Prediction and Risk Factor Analysis Based on a Time–Sensitive, Attentive Neural Network: Retrospective Cohort Study. Journal of Medical Internet Research. 2020;22(7):e16981. Available from: https://www.jmir.org/2020/7/e16981.

10. Lisspers K, Ställberg B, Larsson K, Janson C, Müller M, Łuczko M, et al. Developing a Short–Term Prediction Model for Asthma Exacerbations From Swedish Primary Care Patients’ Data Using Machine Learning — Based on the ARCTIC Study. Respiratory Medicine. 2021;185:106483.

11. Zein JG, Wu CP, Attaway AH, Zhang P, Nazha A. Novel Machine Learning Can Predict Acute Asthma Exacerbation. Chest. 2021;159(5):1747–57.

12. Murugan A, Kandaswamy S, Ray E, Gillespie S, Orenstein E. Effectiveness of a Vendor Predictive Model for the Risk of Pediatric Asthma Exacerbation: A Difference–in–Differences Analysis. Applied Clinical Informatics. 2023;14(5):932–43.

13. Sills MR, Ozkaynak M, Jang H. Predicting Hospitalization of Pediatric Asthma Patients in Emergency Departments Using Machine Learning. International Journal of Medical Informatics. 2021;151:104468.

14. AlSaad R, Malluhi Q, Janahi I, Boughorbel S. Predicting Emergency Department Utilization Among Children With Asthma Using Deep Learning Models. Healthcare Analytics. 2022;2:100050.

15. Hurst JH, Zhao C, Hostetler HP, et al. Environmental and Clinical Data Utility in Pediatric Asthma Exacerbation Risk Prediction Models. BMC Medical Informatics and Decision Making. 2022;22:108.

16. Patel SJ, Chamberlain DB, Chamberlain JM. A Machine Learning Approach to Predicting Need for Hospitalization for Pediatric Asthma Exacerbation at the Time of Emergency Department Triage. Academic Emergency Medicine: Official Journal of the Society for Academic Emergency Medicine. 2018;25(12):1463–70.

17. Chen W, Puttock EJ, Xie F, Crawford W, Schatz M, Vollmer WM, et al. Symptoms of Asthma Extracted Through Natural Language Processing and Their Associations With Acute Asthma Exacerbation in Adults With Mild Asthma. The Journal of Allergy and Clinical Immunology: In Practice. 2025;13(7):1719-29.e7.

18. Xie F, Zeiger R, Saparudin M, Al-Salman S, Puttock E, Crawford W, et al. Identifying Asthma–Related Symptoms From Electronic Health Records Using a Hybrid Natural Language Processing Approach Within a Large Integrated Health Care System: Retrospective Study. JMIR AI. 2025;4:e69132. Available from: https://ai.jmir.org/2025/1/e69132.

19. Afshar M, Press VG, Robison RG, Kho AN, Bandi S, Biswas A, et al. A computable phenotype for asthma case identification in adult and pediatric patients: External validation in the Chicago Area Patient-Outcomes Research Network (CAPriCORN). The Journal of asthma: official journal of the Association for the Care of Asthma. 2018;55(9):1035–42.

20. Fishe J, Pan J, Fedele D, Lyu M, Henson M, Menze N, et al. COMPAC: COMputable Phenotype for Asthma in Children. Research Square. 2025. Preprint.

21. Farion KJ, Wilk S, Michalowski W, O’Sullivan D, Sayyad-Shirabad J. Comparing predictions made by a prediction model, clinical score, and physicians: pediatric asthma exacerbations in the emergency department. Applied Clinical Informatics. 2013;4(3):376–91.

22. Xu M, Tantisira KG, Wu A, Litonjua AA, Chu JH, Himes BE, et al. Genome Wide Association Study to predict severe asthma exacerbations in children using random forests classifiers. BMC Medical Genetics. 2011;12:90.

23. Rezaeiahari M, Brown CC, Eyimina A, Perry TT, Goudie A, Boyd M, et al. Predicting pediatric severe asthma exacerbations: an administrative claims-based predictive model. Journal of Asthma. 2023;61(3):203–11.

24. Overgaard SM, Peterson KJ, Wi CI, Kshatriya BSA, Ohde JW, Brereton T, et al. A Technical Performance Study and Proposed Systematic and Comprehensive Evaluation of an ML-based CDS Solution for Pediatric Asthma. In: AMIA Joint Summits on Translational Science Proceedings; 2022. p. 25–35.

25. Seol HY, Shrestha P, Muth JF, Wi CI, Sohn S, Ryu E, et al. Artificial intelligence-assisted clinical decision support for childhood asthma management: A randomized clinical trial. PLOS ONE. 2021;16(8):e0255261.

26. Denny JC, Ritchie MD, Basford MA, Pulley JM, Bastarache L, Brown-Gentry K, et al. PheWAS: Demonstrating the Feasibility of a Phenome-Wide Scan to Discover Gene–Disease Associations. Bioinformatics. 2010;26(9):1205–10. Available from: 10.1093/bioinformatics/btq126.

27. Yang X, Chen A, PourNejatian N, et al. A Large Language Model for Electronic Health Records. npj Digital Medicine. 2022;5:194. Available from: 10.1038/s41746-022-00742-2.

28. Lundberg SM, Lee S. A Unified Approach to Interpreting Model Predictions. In: Advances in Neural Information Processing Systems 30 (NeurIPS 2017). Curran Associates, Inc.; 2017. p. 4765–74. Available from: https://proceedings.neurips.cc/paper/2017/file/8a20a8621978632d76c43dfd28b67767-Paper.pdf.

29. Licari A, Magri P, Silvestri AD, Giannetti A, Indolfi C, Mori F, et al. Epidemiology of Allergic Rhinitis in Children: A Systematic Review and Meta-Analysis. The Journal of Allergy and Clinical Immunology: In Practice. 2023;11(8):2547–56.

30. Acevedo-Prado A, Seoane-Pillado T, López-Silvarrey-Varela A, et al. Association of rhinitis with asthma prevalence and severity. Scientific Reports. 2022;12:6389.

31. Sheehan WJ, Mauger DT, Paul IM, Moy JN, Boehmer SJ, Szefler SJ, et al. Acetaminophen versus Ibuprofen in Young Children with Mild Persistent Asthma. The New England Journal of Medicine. 2016;375(7):619–30.

32. Baxter L, Cobo MM, Bhatt A, et al. The association between ibuprofen administration in children and the risk of developing or exacerbating asthma: a systematic review and meta-analysis. BMC Pulmonary Medicine. 2024;24:412.

33. Specialist Pharmacy Service (SPS). Using NSAIDs in asthma; 2024. Published 17 July 2024. SPS — The first stop for professional medicines advice. Available from: https://www.sps.nhs.uk/articles/using-nsaids-in-asthma/.

34. Corsico AG, Leonardi S, Licari A, et al. Focus on the cetirizine use in clinical practice: a reappraisal 30 years later. Multidisciplinary Respiratory Medicine. 2019;14:40.

35. Global Initiative for Asthma (GINA). Global Strategy for Asthma Management and Prevention—2025 Update: Guideline Summary; 2025. Publication date: May 6, 2025; last updated: May 7, 2025. Guideline Central. Available from: https://www.guidelinecentral.com/guideline/41774/.

36. National Asthma Education and Prevention Program Coordinating Committee (NAEPPCC) Expert Panel Working Group. 2020 Focused Updates to the Asthma Management Guidelines. Bethesda, MD: National Heart, Lung, and Blood Institute, National Institutes of Health; 2020. NIH Publication No. 20-HL-8140. Available from: https://www.nhlbi.nih.gov/health-topics/asthma-management-guidelines-2020-updates.

